# Dengue Epidemiology in a Brazilian Community

**DOI:** 10.1101/19011965

**Authors:** Wilson Mansho, Fernando Ferreira, Iná Kakitani, Raymundo Soares Azevedo, Marcos Amaku

## Abstract

Dengue is a major vector-borne disease and has motivated health surveillance systems to implement various measures to control it. The epidemiological characterization of dengue occurrence in a community is an important step to foster control activities. We carried out an epidemiological study of the notified and confirmed cases of dengue, from 2000 to 2005, in Guarulhos, State of São Paulo, Brazil. A statistical analysis was performed to test the differences, by sex and age, between the database and the individuals of the population. We also performed a time series analysis of the cases from 2000 to 2007. No statistically significant differences (P>0.05) were observed between the proportions for each sex in the data base and in the city population, and in the categories of 15-19 years, 20-24 years, 25-29 years, 55-59 years, 60-64 years and 70-74 years when compared with the corresponding age intervals in the population. In the other age intervals, statistically significant differences were observed (P<0.05). We observed a cyclic variation in the dengue incidence, between 2000 and 2007, with an alternation of two years with a smaller number of cases (2000-2001 and 2004-2005) and two years with a larger number of cases (2002-2003 and 2006-2007). In the seasonality analysis, the number of cases between February and May was higher than the monthly average. Analyzing the spatial distribution of the cases, we observed the process of increasing endemicity of dengue.

## 1. Introduction

Dengue is the most important vector-borne disease in terms of morbidity, mortality, and economic spending [1] and is even cited in the Book of Records as the world’s largest hemorrhagic fever and vector-borne disease [2]. Dengue has been increasingly worrying countries each year and has led health surveillance systems to adopt various measures to control it.

It is a severe acute febrile illness caused by an Arbovirus of the genus *Flavivirus* of the *Togaviridae* family, of RNA genome and has four serotypes (DEN-1, DEN-2, DEN-3, DEN-4), transmitted by mosquitoes of the genus Aedes and spread mainly in tropical countries [3]. Over 2.5 billion people are at risk of contracting the disease that causes about 100 million cases per year and 100,000 deaths [4].

The economic damage caused by dengue is huge. In Cuba, for example, spending on the 1981 epidemic amounted to $103 million. In addition to direct losses, there are indirect losses, caused by the loss of the workforce of patients and consequent decrease in productivity, losses affecting the tourism sector, absences in schools and the loss of social activities of each patient [5].

In Brazil, the first recorded case of dengue occurred in 1916, in São Paulo, and then in the 1920s, in Rio de Janeiro, and only in 1981, in Roraima, did the disease occur again by serotypes DEN-1 and DEN-4. In 1986, new cases began to occur for the DEN-1 serotype in Rio de Janeiro and later in other Southeast and Northeast states of the country. In 2002, there was a large epidemic in the country, with about 700,000 recorded cases, caused by serotypes DEN-1, DEN-2 and DEN-3 [3].

There are several risk factors for dengue and they interact with each other, further increasing the risk of having the disease in a particular region. The main factors are related to the environment and climate variations, the vector, the target population, and the virus. Dengue is an essentially urban disease, and it is in this environment where the main transmission factors of the disease are found [4]. There are other factors that favor the spread of the vector and the disease, such as the lack of qualified labor for the control service, and the lack of interest of all government levels in investing to reduce the risk factors of the disease [6].

The study of these factors is fundamental for the planning of strategies and the guidance of disease control actions in various aspects, whether in the target population, vector, environment or virus.

The increased incidence of dengue is also related to climate variation, both in temperature and humidity, and even the El Niño phenomenon has been implicated in the increase of cases of various vector-borne diseases [4]. This increase of cases is due to the relationship between climatic factors and vector mosquito behavior. Increasing temperature provides greater distribution and abundance of insects due to the adaptation of the vector to the environment [7]. Temperature is also directly related to period of development of all immature stages. The higher the ambient temperature, the shorter the vector development period and vice versa [8]. Increased rainfall provides the emergence of natural and artificial oviposition sites, as well as the development of mosquito larvae [7]. Altitude is also another limiting variable for vector reproduction, but outbreaks have occurred in places where it was believed that the disease could not occur due to mosquito adaptation and the influence of other risk factors [9].

Regarding gender differences, there are some studies reporting a higher incidence in men, but a greater number of severe cases of dengue hemorrhagic fever (DHF) and dengue shock syndrome (SCD) among women [4]. Other studies found no significant differences for this variable [10].

Regarding age, some studies show that the severity of the disease often affects children and adolescents under 15 years. However, in a study conducted in Puerto Rico, which compared the predisposition of the disease between young and adults, it was observed that older adults were more likely to develop the more severe forms of the disease, similar to children [11].

Another major factor, not only regarding dengue but also contributing to the emergence of other epidemics in various parts of the world, is globalization, which has made trade between countries intensified and increased flow of people between countries [12].

The increase in the disease epidemic worldwide is related to the presence of multiple circulating serotypes in the environment, the rate of genetic change of the virus and the emergence of variants with high epidemic potential, high virulence and importance as a risk factor for dengue hemorrhagic fever (DHF) [5].

In 2004, there were 3 circulating serotypes (DEN-1, DEN-2 and DEN-3) in the state of São Paulo [13].

Dengue virus has been circulating in the city of Guarulhos since the 1990s. This period is the same period that probably led to the definitive installation of *Aedes aegypti* in the state of São Paulo [7].

In this paper, we analyze data from notified cases of dengue in the city of Guarulhos with respect to temporal variations, climatic and socioeconomic variables. We have chosen to analyze the time series from 2000 to 2007 due to the particular pattern of cyclic variation observed in this period.

## 2. Material and Methods

Guarulhos is one of the municipalities of the metropolitan region of São Paulo and is 17 km away from the city of São Paulo. The estimated total population in 2005 was 1,230,511 inhabitants, with 626,514 women and 603,997 men. The demographic density for 2005 was 3684.16 inhabitants per km^2^.

### 2.1. Study design

We used data from notified cases of dengue from the Brazilian Ministry of Health’s Notification Disease Information System (SINAN), installed in the Epidemiological Surveillance Department of the Guarulhos municipality. Importantly, the information in the notification database represents only a part of the total number of infections, because there are infected people who are asymptomatic or have symptoms and were not captured by the municipality’s health system [14].

### 2.2. Analysis of SINAN data

SINAN was created by the National Center for Epidemiology in 1990 to collect and process data on notification of diseases across the country, providing information for morbidity analysis [15]. Data from the SINAN program were collected from January 2000 to October 2005 through notification of suspicious cases by the health units and epidemiological surveillance of the municipality. Also included were cases resident in Guarulhos and reported by other municipalities. The database has been revised to remove duplicate notifications and those with an excess of missing information. Several notification records lacked address information, case closure (open records), and case classification. These records, as well as cases of residents of other municipalities, were not considered in the analysis. The cases were confirmed by serology, by dengue-specific antibody screening, or by the epidemiological link when the epidemic was already in place. The collection of material for serology was careful, each sample was properly packaged and labeled and the material was collected in the correct date (6 days after the onset of the first symptoms). Until 2005, the serology was performed at the Adolfo Lutz Institute and from that year began to be performed at the Public Health Laboratory of Guarulhos. All reported cases were assessed according to the suspicious case definition criteria used by the Ministry of Health: abrupt onset high fever lasting up to 7 days, accompanied by at least two of the following symptoms: headache, retroorbital pain, myalgia, arthralgia, prostration and rash [3].

From the SINAN database, some socio-demographic variables (gender and age) and variables related to signs and symptoms of each case were selected.

For the variable “gender”, a comparison of proportions was performed using the Minitab 14 program (Minitab Inc., 2004) for a significance level of 5%. The proportions of the total population that served as reference for the Guarulhos female and male populations were, respectively, 50.9% and 49.1%. To obtain these values, the Guarulhos population average was calculated for the period 2000-2005 (female population average of 584,299 and male population average of 564,258), based on data from SEADE (2006) [16].

The proportion of cases in each age group was compared to the population proportion of that age group in SEADE data, similar to the comparison performed for gender. The average of each age category for the study period was also calculated.

The seasonal index was calculated in the R 3.6.1 program [17] using the time series decomposition method with a multiplicative model [18]. To perform this calculation, data from confirmed and notified cases of SINAN from 2006 and 2007 distributed in each month of the year were included. In addition to the seasonal index, an estimate of the temporal trend was performed and the presence of a cyclic component was verified.

### 2.3. Analysis of meteorological data

Rainfall and temperature data for the years 2000-2005 were provided by the National Institute of Meteorology (INMET). A simple linear regression analysis was performed for 2002 and 2003, years related to the peak of the epidemic in the municipality.

### 2.4. Georeferenced maps and case distribution on maps

The georeferenced maps were provided by the Geoprocessing Sector of the Department of Informatics and Telecommunications of the Secretariat of Administration and Modernization of Guarulhos. For some cases, there was only information regarding the neighborhood of residence, without full address. The cases were then distributed on the map across the city’s neighborhoods and can be observed by color gradient.

Dengue cases were mapped and their distribution in the municipality was analyzed using the ArcView GIS 9.1 program (ESRI Inc., 2005).

## 3. Results

Of the 774 cases reported, 409 were female (52.8%, 95% CI: [49.3%; 56.4%]) and 365 male (47.2%, 95% CI: [43.6 %; 50.1%]). When we applied the two-proportion comparison test to compare the proportion of cases in women with the proportion of women in the population (based on 2005 data), we did not observe statistically significant differences (P = 0.28).

Although a large proportion of the population of the city is in the age group from 1 to 24 years old (47.6%), most cases belong to the age group of 20 to 44 years old (53.9%). The distribution of cases by age group can be seen in **Figure 1**. We compared, by age group, the proportion of dengue cases and the proportion of individuals in the population in that age group (mean from 2000 to 2005). The results can be seen in **Figure 1**. We observed that in the age groups of 15-19 years, 20-24 years, 25-29 years, 55-59 years, 60-64 years and 70-74 years there was no statistically significant difference between the proportions (P> 0.05). In the other age groups, on the other hand, significant statistical differences were observed between the proportion of dengue cases and the proportion of individuals in the population in that age group (P<0.05).

**Figure 1.**
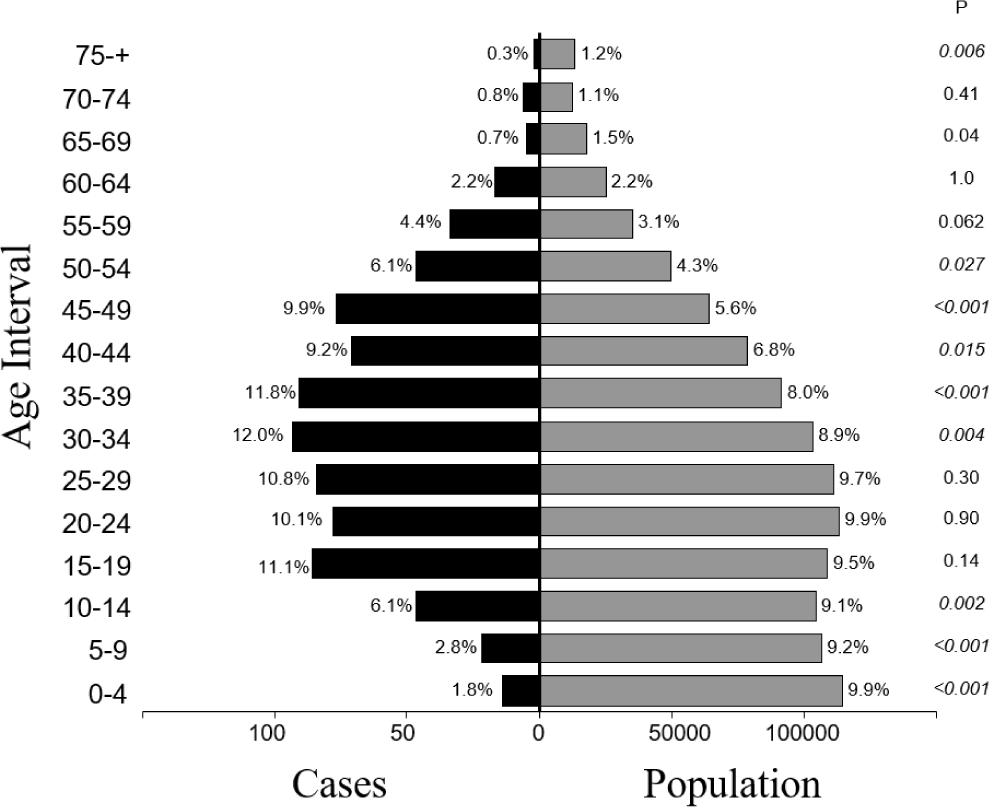
Distribution of confirmed cases of dengue and population by age group, in absolute and relative numbers, in Guarulhos, from 2000 to 2005. The column on the right indicates the P values of the comparison of proportions test. P values less than 0.05 are in italics.

The most prevalent symptoms reported by the notified and confirmed cases, in addition to fever, were headache with 692 cases (89.4%, 95% CI: [87.0%, 91.2%]), myalgia with 668 cases (86.3%, 95% CI: [83.7%, 88.7%]), prostration with 576 cases (74.4%, 95% CI: [71.1%, 77.5%]) and arthralgia with 541 cases (69.9%, 95% CI: [66.5%, 73.1%]). The frequency of symptoms can be seen in **Table 1**. About 312 cases (40.3%, 95% CI: [39.0%, 46.1%]) had rash and 53 cases (6.8% 95% CI: [5.2%, 8.9%]) had petechiae, 42 cases had shock and 129 were hospitalized. The most frequent symptoms varied over the years and some symptoms were observed in some years and not observed in other years. There was also a variation in the severity of the disease, which can be observed by a higher frequency of hemorrhagic phenomena and shock in some years and also by the number of hospitalizations that occurred during the study period. Most cases evolved to cure, except in 2 of them who died.

**Table 1.** Frequency of signs and symptoms of reported and confirmed cases of dengue in the city of Guarulhos, from 2000 to 2005.

| Signs / Symptoms | Frequency (%) |
| --- | --- |
| headache | 89.4 |
| myalgia | 86.3 |
| prostration | 74.4 |
| arthralgia | 69.9 |
| retro-orbital pain | 69.1 |
| nausea | 65.5 |
| rash | 40.3 |
| diarrhea | 27.8 |
| abdominal pain | 26.2 |
| petechiae | 6.8 |
| epistaxis | 5.7 |
| shock | 5.4 |
| gingivorrhagia | 3.0 |
| hypotension | 2.8 |
| bleeding | 1.2 |
| hepatomegaly | 1.2 |
| metrorrhagia | 0.8 |
| ascites | 0.8 |

Although tourniquet test is considered essential for the diagnosis of DHF [19] [20], it was performed only in 98 notified and confirmed patients. If we count all 2864 notifications made from 2000 to 2005, this test was performed only on 337 notifications.

### 3.1. Analysis of seasonal index and temporal trend

**Figure 2** shows the seasonal index calculated from monthly dengue incidence data from January 2000 to December 2007. Months in which the average frequency of cases is higher than the annual average (calculated from monthly values) have a seasonal index above 1; in addition, months with an average frequency below the annual average have a seasonal index below 1. We note that the seasonal index is higher than the annual average between February and May.

**Figure 2.**
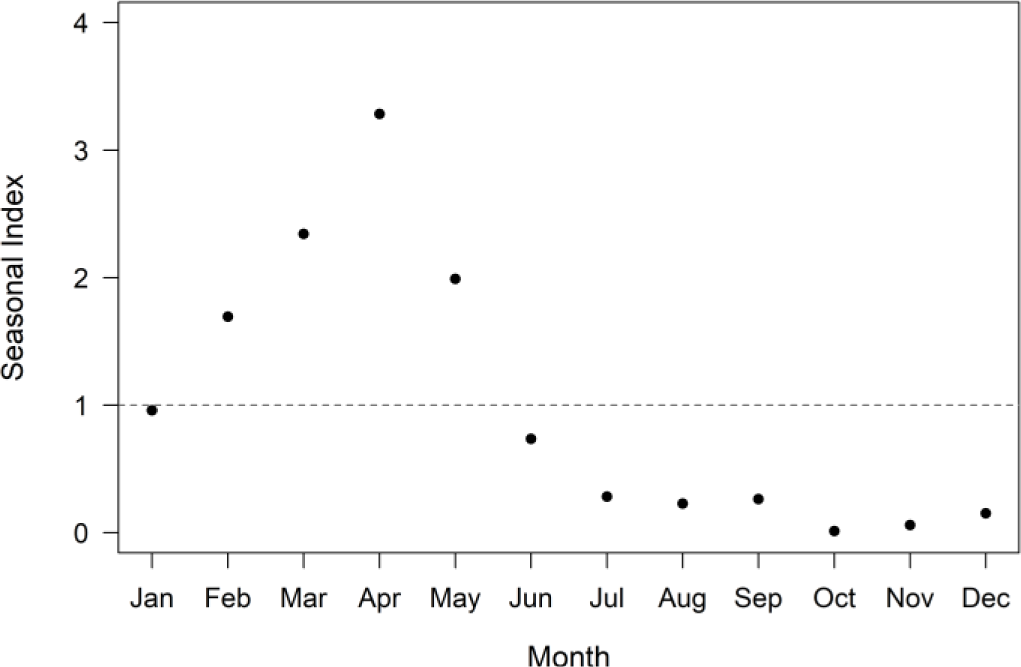
Seasonal index of dengue cases during the months of the year from January 2000 to December 2007, in the city of Guarulhos.

The temporal trend analysis showed an increasing trend (**Figure 3**). In this figure it is possible to observe the process of endemization, with the occurrence of cases every year. We also note the alternation of two years with a smaller number of cases (2000-2001 and 2004-2005) and two years with a comparatively larger number of cases (2002-2003 and 2006-2007).

**Figure 3.**
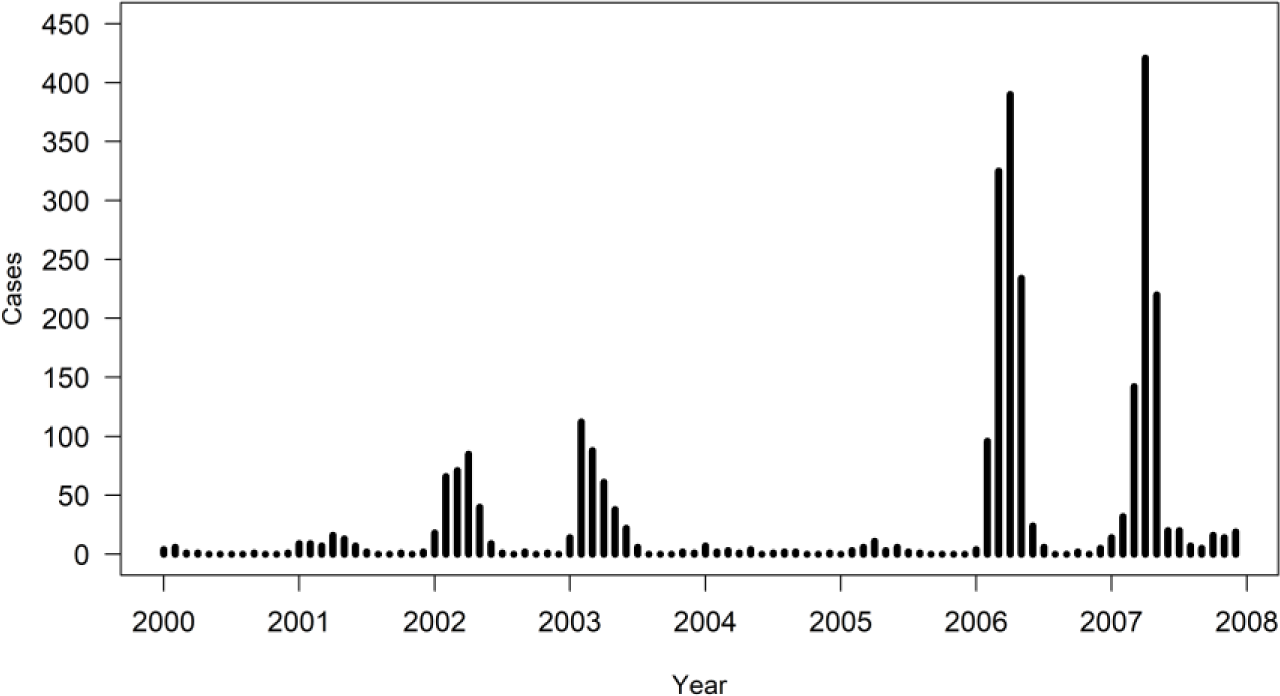
Time series of confirmed cases of dengue by month from January 2000 to April 2007 in Guarulhos.

### 3.2. Linear regression analysis of reported cases with regard to monthly rainfall and average temperature

The simple linear regression of the notified and confirmed cases with regard to rainfall and average temperature did not show any relationship between these variables. For the monthly total precipitation and the average temperature, we tested the models with the data from 2000 to 2005, from 2001 to 2003 and only the data from 2002 and 2003.

**Figure 4** shows, as an example, the number of reported cases, the average temperature and the monthly rainfall for 2003. We note that the rainfall decreased from January to June and increased from July to December. The lowest average temperatures were observed between May and August. The seasonal index (**Figure 2**) reflects the pattern observed in the number of cases.

**Figure 4.**
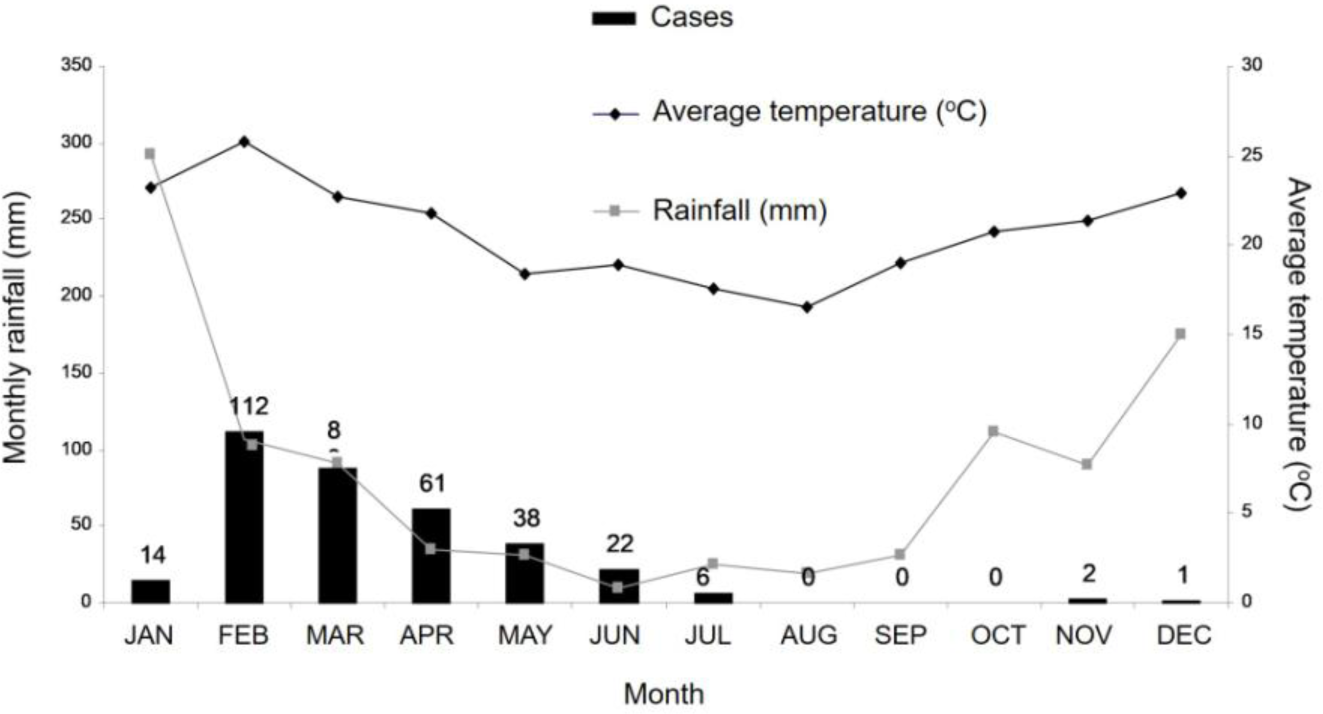
Cases of dengue, average monthly temperature (ºC) and monthly rainfall in 2003 by month in Guarulhos.

**Figure 5**, referring to 2003, exemplifies the spatial distribution of the notified and confirmed cases in Guarulhos. We noticed that the cases were spread all over the city except for a few neighborhoods.

**Figure 5.**
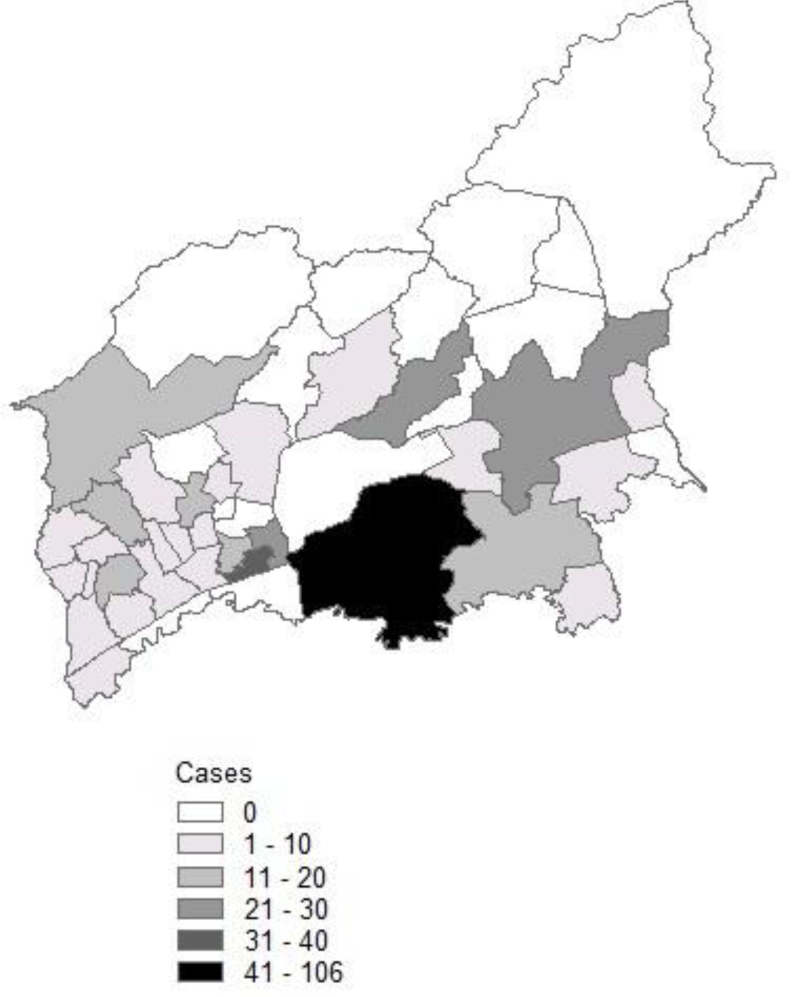
Spatial distribution of confirmed cases of dengue in Guarulhos in 2003.

## 4. Discussion

The two-proportion comparison test applied to the gender variable indicates that the disease did not affect both genders differently. These differences were also not found in other research, such as seroepidemiological surveys conducted in Brazil [10]. In research conducted in other countries, a significant difference was observed between the genders [4].

Most of the cases occurred in age groups of great importance in the city economy, causing losses due to absenteeism. Although there are few cases in adults older than 65 years (1.7%) and in children 0-14 years (10.7%), these age groups should be monitored and appropriate treatment provided, since the risk of developing the most severe forms is much higher than in other age groups [11] [21]. Burattini et al. (2016) [22] observed that the risk of hospitalization for dengue in Brazil is higher for children aged less than 10 years and for adults older than 65 years.

Although no statistical correlation was observed between the number of cases each year and the variables of monthly precipitation and average temperature, we cannot disregard this hypothesis, since there are several studies that established a strong relationship between the incidence of dengue and rainy seasons, high temperatures, altitudes and winds [9]. On the other hand, the seasonal index analysis showed that there is a concentration of reported cases from February to May.

Observing the graph of dengue cases, average temperature and rainfall distributed by months for 2003 (**Figure 4**), for example, we note that the number of cases decreased from February to June, in line with decreases in average temperature and total rainfall. In the other months, however, the low number or absence of cases does not allow a relationship between these variables to be observed. The low correlation observed between the variables may be related to months without cases and/or to the time lag between the increase of temperature and rainfall during the spring and early summer, the subsequent increase of the mosquito population and the occurrence of dengue cases in the human population. In addition, we note that the number of cases exceeds the annual average from February to May and is below average from June to January.

An aspect that caught our attention was the cyclic variation in the dengue incidence from 2000 to 2007, with a two-year alternation with a smaller number of cases (2000-2001 and 2004-2005) and two years with a comparatively larger number of cases (2002-2003 and 2006-2007). This pattern of outbreaks could be understood as the result of the spread of dengue across a range of neighborhoods within the city, associated with the proportion of the remaining susceptible individuals in each neighborhood and the human movement between neighborhoods, as described by Amaku et al. (2016) [23] using mathematical modeling.

Through the analysis of the time series, it is possible to observe the process of endemization, with the occurrence of cases every year, which was also observed in other works [14].

Looking at the georeferenced map for 2003 (**Figure 5**), it is also possible to observe the process of disease endemization, with cases spread throughout the city. We can also, through the use of geographic information systems, plan strategies and improve entomological and epidemiological surveillance activities to prevent the spread of the disease in places with high transmission potential. Geographic information systems are already being used by several municipalities for dengue control allowing for a joint analysis of the data [24].

Regarding the limitation of the study, maps could be used more precise and accurately if the full standardized address database was available in SINAN. This problem was also reported by Skaba et al. (2004) [25]. Several notification records lacked full address information, thus it was not possible to perform a more detailed spatial analysis. Information on the dengue serotype that caused infection in each patient was also limited.

Based on the results obtained, we can consider some points for the improvement of dengue surveillance and prevention actions, such as the use of maps for the planning of vector control areas, the monitoring of the temporal trend of cases, and the analysis of the incidence of cases according to socio-demographic variables, to observe if there is any trend of higher incidence in any specific category.

## 5. Conclusion

We observed a cyclic variation in the dengue incidence in Guarulhos, between 2000 and 2007, with an alternation of two years with a smaller number of cases (2000-2001 and 2004-2005) and two years with a larger number of cases (2002-2003 and 2006-2007). In the seasonality analysis, the number of cases between February and May was higher than the monthly average. Dengue affected both genders similarly. Most of the cases occurred in age groups of great importance in the city economy, causing losses due to absenteeism. Analyzing the spatial distribution of the cases, we observed the process of dengue endemization.

## Data Availability

The dataset of dengue cases is attached as supplementary material.

## Acknowledgements

We thank Regina Laudari and Cristina Magnabosco for their support and data provided by the Municipality of Guarulhos; the professionals of the Geoprocessing Sector of Guarulhos for providing the georeferenced maps and the address database; and Alaor Moacyr Dall’Antonia Junior and Cristina Costa for data provided by the National Institute of Meteorology (INMET). The authors acknowledge financial support from CNPq. This study was financed in part by the Coordenação de Aperfeiçoamento de Pessoal de Nível Superior - Brasil (CAPES) - Finance Code 001.

